# Improving Numerical Measures of Human Feelings: The Case of Pain

**DOI:** 10.1101/2025.03.05.25323444

**Authors:** Michele Garagnani, Petra Schweinhardt, Philippe N. Tobler, Carlos Alós-Ferrer

## Abstract

Numerical self-report scales are extensively used in economics, psychology, and even medicine to quantify subjective feelings, ranging from life satisfaction to the experience of pain. These scales are often criticized for lacking an objective foundation, and defended on the grounds of empirical performance. We focus on the case of pain measurement, where existing self-reported measures are the workhorse but known to be inaccurate and difficult to compare across individuals. We provide a new measure, inspired by standard economic elicitation methods, that quantifies the negative value of acute pain in monetary terms, making it comparable across individuals. In three preregistered studies, 330 healthy participants were randomly allocated to receive either only a high- or only a low-pain stimulus or a high-pain stimulus after having double-blindly received a topical analgesic or a placebo. In all three studies, the new measure greatly outperformed the existing self-report scales at distinguishing whether participants were in the more or the less painful condition, as confirmed by effect sizes, Bayesian factor analysis, and regression-based predictions.

## 1 Introduction

The study of self-reported, subjective feelings has been cited as one of the fastest growing research fields in economics over the last two decades (e.g., Kahneman and Krueger, 2006; Stutzer and Frey, 2012; Chen et al., 2022; Liu and Netzer, 2023). Self-reported numerical measures of human feelings are extensively used to assess subjective variables in economics, psychology, and medicine, and are routinely collected by national and international organizations in large-scale panels (e.g., Office for National Statistics, 2023). These scales quantify a variety of feelings, from discrete emotions to life and job satisfaction, individual happiness, and the subjective experience of pain. However, such scales are often criticized because they lack a scientific, objective foundation. For example, many economists have argued that, since the numbers used in subjective scales do not correspond to objective measurements and cannot be compared across individuals, the results of those scales are essentially meaningless (Bond and Lang, 2019). In contrast, Kaiser and Oswald (2022) have argued that numerical (even integer) scales of human feelings are empirically informative because they often exhibit a robust correlation with subsequent actions (e.g., moving houses, partner separation, changing jobs) and a greater predictive power than more objective, socioeconomic variables (e.g., age, marital status, education).

Kaiser and Oswald (2022) conclude that, in some cases, human beings seem to be able to successfully operationalize abstract numerical scales to measure the intensity of feelings even when no absolute, objective measurement exists. However, this might not always be the case. A prominent example where self-reported numerical scales are extensively used in practice is the measurement of pain intensity. The gold standard in this field is given by self-reported measures relying on visual, numerical, or verbal one-dimensional rating scales (Wallenstein et al., 1980), which are variations of questions such as “on a scale of zero to ten, how much does it hurt?” Numerical scales like these have been criticized as inaccurate (Huskisson, 1974; Breivik et al., 2008; Phillips, 2009; Von Korff et al., 2020), are very difficult to compare across people (McGuire, 1984; Jensen et al., 1986; Coll et al., 2004), and often produce inconsistent and biased scores (Robinson-Papp et al., 2015; Ackerman et al., 2020). Illustrating the inconsistency across different measurement methods, estimates of the prevalence of chronic pain in adults range from 18% to 34.5% for the general population (Dahlhamer, 2018; Pitcher et al., 2019), and from 1.4% to 41.6% for young adults (Murray et al., 2022). Thus, better measurements are needed.

Pain measurements are one of the most socially relevant applications of subjective, numerical scales. It is widely accepted that inaccurate pain measurements can lead to inadequate pain management and a reduction in quality of life (Phillips, 2009). They can also impose a considerable burden on health care systems. For example, more than US $600 billion are spent annually in the USA to treat pain, surpassing the cost of treating heart disease and diabetes (Gaskin and Richard, 2012). Inaccurate pain measurement might also be an important factor contributing to the opioid epidemic in the United States (Stern and Roberts, 2016; McGreal, 2018). Accurate and sensitive measurements of pain also matter for medical practice, because pain cannot be considered relieved unless it has been measured (Huskisson, 1974).

In this contribution, we focus on the measurement of subjective pain and, in particular, the discriminability of different intensities of pain stimulation across individuals. We aim to contribute to the debate on the relevance of numerical scales of human feelings while concentrating on a particular but economically consequential application. Specifically, we propose and empirically validate a new measure of acute, experimentally-induced pain across individuals. In three preregistered randomized controlled trials, we show that the new measure greatly outperforms established procedures at differentiating human participants according to which acute pain levels they experienced. That is, our studies focus on the *signal extraction problem*, where the experimenter randomly assigns participants to two groups who experience objectively-different painful stimuli, and evaluates scales on their ability to statistically differentiate the groups. The measurement of experienced pain across (groups of) individuals is particularly important for clinical studies with random parallel assignment of participants to groups, e.g. as required to evaluate the effectiveness of pain-relief treatments.

The idea behind the new measure is to provide empirical content for numerical quantities in a way that facilitates interpersonal comparisons. Our scale provides a bridge between subjective, incomparable intensities and objective, interpersonally comparable magnitudes by relying on monetary quantities, which are familiar to all individuals. Specifically, the scale measures how much money a person would be willing to accept in order to re-experience a painful stimulus. While such a comparison can also be criticized on theoretical grounds (as is the case of abstract, integer measures), we adopt an empirical approach for performance evaluation (as Kaiser and Oswald, 2022). The intuition behind our approach is simply that familiar monetary quantities will make it easier for individuals to operationalize the scale in a way that more strongly correlates with other individuals’ evaluations.

The logic of the experimental studies and the validation of the proposed method is as follows. Subjective pain in response to a nociceptive stimulus (i.e., a stimulus which induces the perception of pain) of a fixed intensity varies across individuals, and hence any pain measurement method generates a distribution of measurements for any given stimulus or treatment. To compare the capacity of different pain measures to separate distributions with different means, we randomly assign participants to conditions differing in the objective intensities of the nociceptive stimuli or pain treatment. A better measure of experienced pain should more easily differentiate between conditions, i.e., between participants with high or low intensity of nociceptive stimulation. That is, even though individual measurements for the same painful stimulus will always differ across individuals, alternative measures will vary in their ability and accuracy to statistically detect differences in the distributions arising in low vs. high pain conditions.

We compared two versions of the new method (which we call ME1 and ME2) to the three most-commonly used standard pain scales (Numerical Rating Scale, Visual Analog Scale, and General Labelled Magnitude Scale). For each scale, the distribution of numerical values across individuals in a group quantifies how the experience of pain for an isointense stimulus is distributed among participants. This maps physical intensities to numerical amounts corresponding to the acute pain induced by the reference stimulus. Each pain measurement method generates one distribution of measurements per condition. Thus, for each scale, we compare the distributions for two different intensities of nociceptive stimulation. We then compare scales according to how well they separate the two groups using effect sizes, Bayesian factor analysis, and regression-based predictions with the condition as dependent (predicted) variable. In all three trials, we find that the proposed method greatly outperforms established procedures at differentiating acute pain levels across participants.

## 2 Materials and Methods

### 2.1 Study design

We conducted three separate preregistered randomized controlled experiments (total 330 participants) in the Laboratory for Social and Neural Systems Research (SNS) at the University Hospital of Zurich (Switzerland). In experiments 1 and 2, we varied the pain intensity experienced by participants using two different electrical stimuli (high vs. low voltage; *N* = 109) and contact-heat stimuli (higher vs. lower temperature; *N* = 115), respectively. In the third, we fixed the intensity of the painful stimulus (higher temperature), and tested the method’s ability to differentiate a topical analgesic from a placebo in a double-blind randomized trial (*N* = 106). This approach emulated standard procedures for the evaluation of pain-relief treatments.

In each study, participants were randomly assigned to one of the two groups. All measurements were individual, i.e. there was only one participant present during measurement. Participants were seated in front of a computer screen and received one painful stimulus on the left volar forearm. They then evaluated the painful stimulus according to two versions of the newly-proposed measure, and to the three most commonly-used standard procedures (Figure 2). These are the Numerical Rating Scale (NRS, Fruhstorfer et al., 1976), on which pain intensity is assessed in integers between 0 and 10, the Visual Analog Scale (VAS, Breivik et al., 2008), on which patients mark their pain level on a line of a predefined length without verbal labels, and the General Labelled Magnitude Scale (gLMS, Morin and Bushnell, 1998), on which patients mark their pain level on a line with verbal labels describing the intensity, e.g. Weak or Strong.

In the newly-proposed method participants are repeatedly asked to choose between an amount of money *X* (varying across questions) together with a reference painful stimulus previously experienced at the beginning of the experiment, or a fixed, smaller amount of money but no pain (e.g., *X* = $ 15 and additional pain vs. $ 10 but no pain; see Figure 1). The question where participants switch from choosing the smaller monetary amount to choosing the painful stimulus and the larger monetary amount captures the subjective value of the painful stimulus in monetary terms, i.e., the monetary equivalent (ME) of the reference pain. For example, if a participant chose $ 10 and no pain rather than pain and monetary amounts of $ 12.5, $ 15, or $ 17.5, but chose $ 20 and pain rather than $ 10 and no pain, the ME is calculated as $ 20 *−* 10 = $ 10. The new method was administered once as a list of questions (ME1), ordered according to monetary amount (*X*) combined with the painful stimulus (Figure 1a), and once as a series of sequential binary choices in randomized order (ME2; Figure 1b). ME1 and ME2 are calculated in the same way.

**Figure 1:**
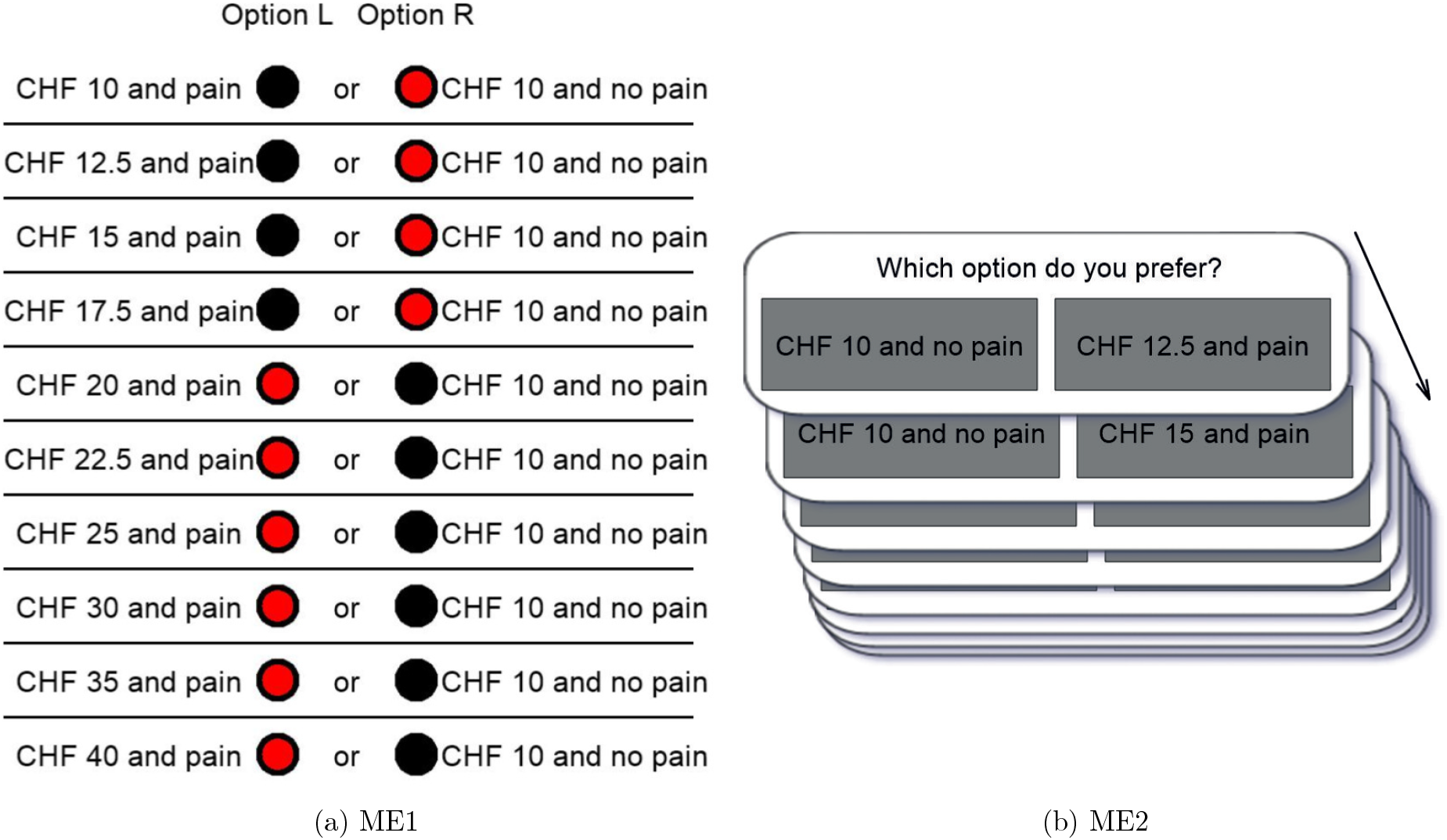
The two implementations of the newly proposed method. (a) ME1 uses a list of questions and enforces consistency. (b) ME2 presents questions in random order and does not enforce consistency (one question per screen). Monetary amounts were in Swiss Francs (CHF).

**Figure 2:**
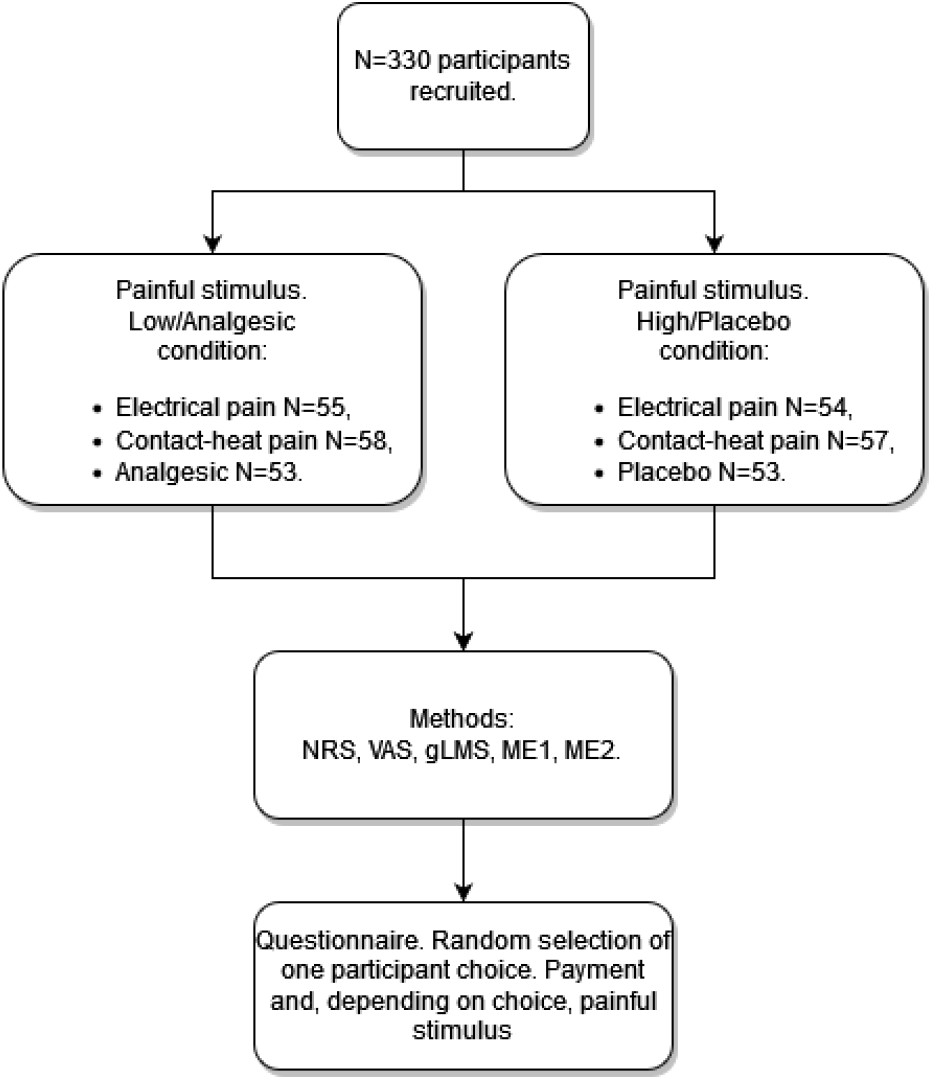
Design of the study. Numerical Rating Scale (NRS), Visual Analogue Scale (VAS), General Labelled Magnitude Scale (gLMS), and the newly proposed method as a list of questions (ME1) or as a series of sequential binary choices in randomized order (ME2)

Our procedure provides a truthful, incentive-compatible numerical measure of how much a treatment (e.g., an analgesic) effectively decreases acute pain intensity, in a way that is comparable across people. The procedure builds upon *multiple price lists* as frequently used in economics for the purpose of truthfully eliciting preferences. These lists determine monetary amounts which leave people indifferent between obtaining an item (such as a lottery: Holt and Laury, 2002, 2005) and receiving the monetary compensation, i.e. the Willingness to Pay or Willingness to Accept (Plott and Zeiler, 2005). Monetary valuations, implemented as multiple price lists or (essentially equivalent) second-price auctions, are also often used to estimate individual and societal preferences in the field (Bateman et al., 2002). In the context of pain, monetary valuations were previously used by Vlaev et al. (2009) to estimate the financial value of pain relief. The only condition for monetary valuation methods to be incentive-compatible is that choosing the subjectively better option results in a larger amount of money than any other choice. In our context, this is guaranteed as long as individuals desire to avoid pain and prefer more money to less.

The order of the methods was fixed (from simpler to more complex) to avoid carryover effects. For example, in the gLMS numbers are associated to labels, while in the NRS numbers have no such association. Hence presenting the gLMS first could create an association, confounding the NRS data.

All monetary amounts used in ME1 and ME2 were stated in Swiss Francs (CHF). In ME1, all binary questions were listed simultaneously, and consistency was enforced (Figure 1a). In ME2, each choice was presented sequentially on the screen (Figure 1b) and participants could be inconsistent, e.g., select the painful stimulus for CHF 15 but no pain for CHF 17.5. In ME2 we used the total number of choices of the smaller monetary amount (and no pain) to determine the individual’s switching point, as is common practice in multiple price lists (Holt and Laury, 2002, 2005; see, however, Chew et al., 2022). Inconsistencies in ME2 occurred for 13 (11.93%), 3 (2.61%), and 5 (4.72%) participants in the electrical pain, heat pain, and analgesic studies, respectively. In ME1 and ME2, participants could choose to receive a painful stimulus for CHF 10 over receiving CHF 10 and no pain. For ME1 this happened for 1 (0.92%), 1 (0.87%), and 0 (0.00%) participants, in the electrical pain, heat pain, and analgesic studies, respectively, while for ME2 it occurred in 4 (3.60%), 2 (1.74%), and 0 (0.00%) participants. Thus, participants in all three studies paid attention and understood the task.

After the main task, participants answered a questionnaire (age, gender, etc.). At the end of the experiment, one of the participant’s choices in the new method was randomly selected by the computer and implemented (also randomly selecting ME1 or ME2). Participants were informed about this at the beginning of the study. That is, participants received what they had chosen in the selected round, either a smaller amount of money and no pain or a larger amount and the painful stimulus. This avoided deception and increased credibility of the measurement.

The studies were approved by the Cantonal Ethics Committee in Zurich (BASEC: 2022-00236) and were conducted in accordance with Swiss law and the Declaration of Helsinki. They were registered with ClinicalTrials.gov, *NCT* 05415423. and with AsPredicted.org (study numbers 98339, 102710, and 104958, respectively).

### 2.2 Participants

Inclusion criteria were 18-60 years of age, willingness to participate in the study, written informed consent, and good English language skills. Exclusion criteria were inability to give informed consent, any neurological disorders, reduced general health or chronic diseases (e.g. autoimmune disease, severe cardiovascular diseases, insulin-dependent diabetes), severe depression, chronic pain, pregnancy, or intake of drugs potentially influencing pain perception in the week before the experimental session (e.g. cannabis). Participants were made aware of these criteria and the topic of the study when they were recruited and reminded on the day of the study. Participants provided written consent before starting the study, and could withdraw from the study at any time without giving a reason.

### 2.3 Randomization and masking

Participants were randomly assigned to a High pain or a Low pain condition. Participants were aware of the existence of these two conditions, but they did not know which one they belonged to. Within each condition participants experienced a physically-identical stimulus. The characteristics of the participants are summarized in Table 1.

**Table 1:** Characteristics of the participants in the three studies by condition. Entries in brackets are SDs, percentages (for gender), or median (for income range).

|  | Electrical pain |  | Contact-heat pain |  | Analgesic |  |
| --- | --- | --- | --- | --- | --- | --- |
|  | Low | High | Low | High | Analgesic | Placebo |
| *N* | 55 | 54 | 58 | 57 | 53 | 53 |
| Female | 26 (47.3%) | 22 (40.7%) | 24 (42.1%) | 25 (43.9%) | 33 (63.5%) | 35 (66.0%) |
| Age (years) | 24.60 (4.43) | 25.43 (4.57) | 24.26 (3.74) | 25.58 (4.12) | 24.60 (2.98) | 25.81 (5.77) |
| Income (in CHF) | 56k (98k) | 80k (79k) | 13k (26k) | 11k (29k) | 90k (164k) | 75k (110k) |
| Range (median) | 0-300k (12k) | 0-500k (70k) | 0-150k (0k) | 0-120k (0k) | 0-600k (38k) | 0-1000k (30k) |
| % with zero income | 18.18% | 33.33% | 61.40% | 58.18% | 30.77% | 22.64% |
| Money valuation | 25.60 (5.00) | 26.37 (3.99) | 26.43 (4.61) | 27.70 (3.93) | 24.70 (4.14) | 27.30 (3.71) |
| Pain Threshold (in °C) |  |  | 45.35 (3.72) | 44.93 (4.16) |  |  |

In the contact-heat study, there was no difference in the pain threshold (measured according to the method of limits; Yarnitsky et al., 1995) between the participants of the two conditions (Low 45.35 °C, SD 3.72, High 44.93°C, SD 4.16; MWU, *z* = 0.295, *p* = 0.7702). Consequently, the pain threshold did not explain individual differences in pain responses (Spearman’s correlation *ρ* = 0.0183 between ME1 and the pain threshold, n.s.).

In the analgesic study, the two groups experienced the same physical pain stimulus after administration of either a placebo (High pain condition) or an analgesic cream (Low pain condition), again randomly assigned. The two creams were indistinguishable and neither the participant nor the experimenter knew which cream was which (double-blind design). The pharmacy which produced the creams was also responsible for the randomization. Participants could not guess the condition they were in. According to the final questionnaire, 95.83% of participants thought they received the placebo, attesting to the success of the blinding procedure and indicating that participants in the analgesic condition also experienced pain.

### 2.4 Procedures

In the electrical pain study, participants experienced electrical stimuli with 10 pulses of 15 ms duration spaced 10 ms apart, administered with a DS5 Isolated Bipolar Constant Current Stimulator (Digitimer), a standard device used in clinical trials to induce pain through electrical pulses. The stimuli had an intensity of 50 mA and a voltage of 10 V in the High condition vs. 5 V in the Low condition. In the contact heat study, participants experienced a 5-second stimulus of 50°C in the High condition or a 48°C stimulus in the Low condition. We used the TSA II from Medoc Ltd, a standard device used in clinical trials to induce pain through contact heat. The temperatures were chosen to prevent any skin damage.

The differences between high and low intensity were determined on the basis of pretests, similarity to previous studies (Jepma et al., 2018), and the need to ensure that the expected participants’ decisions would yield an average remuneration in line with lab-enforced standards. The aim was to use two objectively different temperature levels that would induce clearly separable pain experiences within-subject, and test how well the measures can distinguish them between-subject.

The analgesic study also used the 5-second contact heat stimulus at 50°C. The analgesic was a topical cream (5g; main ingredients Lidocaine base 0.23 g, tetracaine base 0.04 g, tetracaine HCl 0.04 g and miglyol, paraffin oil, high-pressure polyethylene at 1g) and the placebo cream (5g E171, miglyol, paraffin oil, high-pressure polyethylene) was indistinguishable from the verum. They were produced by a pharmacy (Dr.Hysek AG) that holds a manufacturing authorization from Swissmedic in accordance with Art.5 of the Therapeutic Products Act (HMG, *SR*812.21). The production followed Good Manufacturing Practice guidelines regarding compliant manufacture, packaging, randomization, blinding and labeling of medicinal products for clinical trials. After the application on the participants’ volar forearm in an area of 3*×*4 cm, the cream was covered by a foil (OPSITE Flexifix) for 30 minutes to enhance transdermal penetration. The cream was then removed and the painful stimulus applied to the same location.

### 2.5 Preregistration and sample size

All analyses and hypotheses were preregistered (with the exception of additional, exploratory regression analyses) in AsPredicted.org, study numbers 98339, 102710, and 104958, respectively. We did not deviate from the preregistration and we report all the measured variables. Sample sizes were calculated to have a power of 0.8 to detect a medium effect size (*d* = 0.5) difference between the two groups for a non-parametric, two-sided, between-subject test (Mann Whitney U-Test; MWU) with an *α* = 0.05, yielding *N* = 106 participants. We collected *N* = 109, *N* = 115, and *N* = 106 participants for the electrical pain, contact heat pain, and analgesic studies, respectively. All participants were included in the analysis, performed using STATA 16.

## 3 Results

De-identified participants’ data and the code for the analyses is publicly available in OSF (https://osf.io/jtv8p). In total, 330 participants were recruited for the three studies. The expectation was that the High pain condition group should provide higher pain ratings than the Low pain condition group. The new measures (ME1 and ME2) were clearly more accurate and sensitive than the standard procedures (NRS, VAS, and gLMS) in capturing the difference in pain stimulation between the conditions (Figure 3). No participant refused the highest monetary amount, hence it was always possible to identify an ME (of at most 40). This agrees with previous literature showing the existence of money-pain tradeoffs in lab contexts (Vlaev et al., 2009; Slimani et al., 2021).

**Figure 3:**
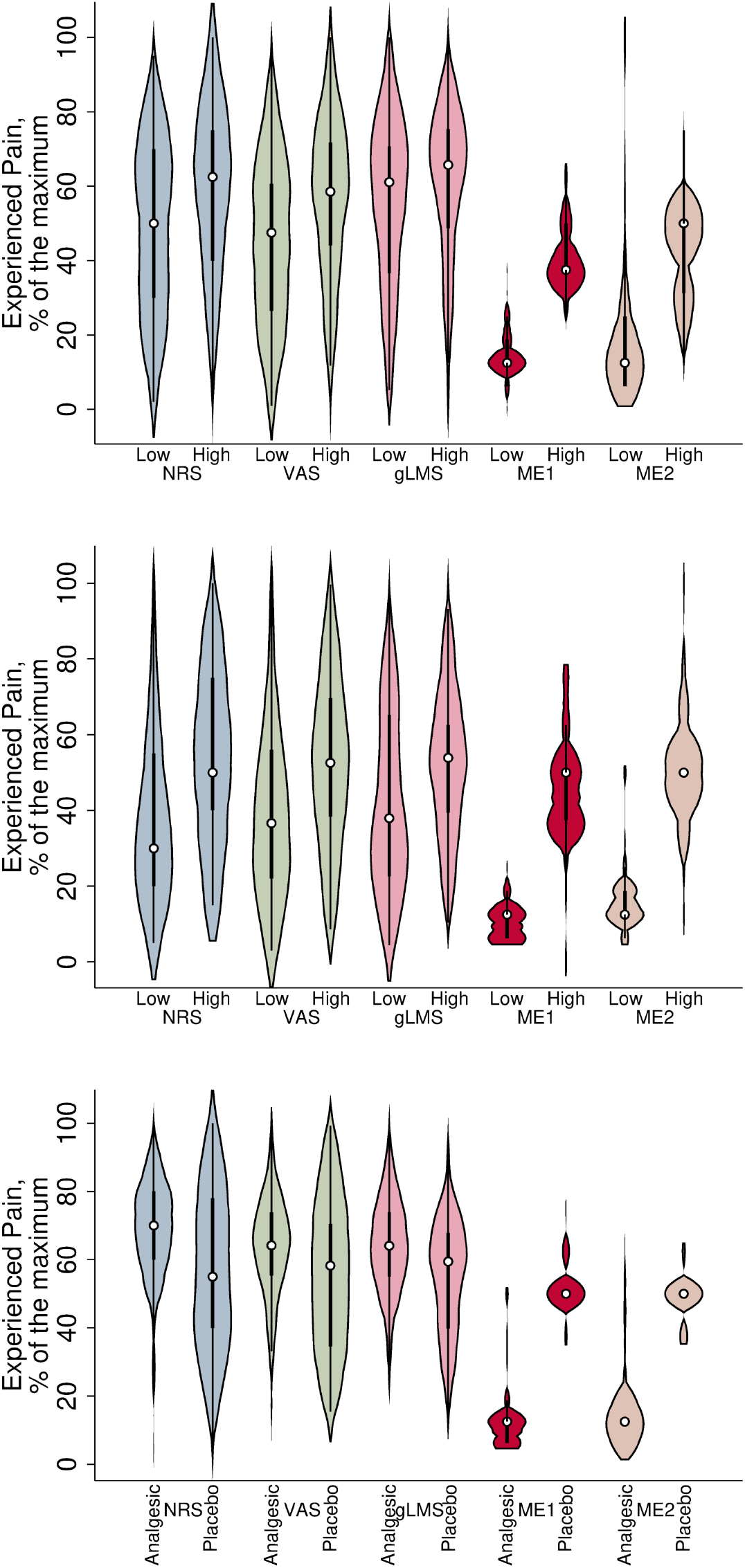
Distribution of reported pain levels as measured by the different methods, split by condition (Low Pain/Analgesic vs. High Pain/Placebo). Scores were normalized to lie between 0 and 100. Violin plots show the median, the interquartile range and the 95% confidence intervals, as well as rotated kernel density plots on each side.

### 3.1 Statistical separation of experimental groups

For the electrical pain study, the differences in pain ratings were significant in the expected direction both for the standard methods and for the two new methods (Mann Whitney U-tests; see Table 2 for all the tests and see Tables A.1 to A.3 in the Appendix for min, max, mean, and SE for each method, condition, and study). Importantly, the separation was more pronounced for the new methods than for the standard ones. The effect sizes of the subjectively-experienced pain induced by the objective difference in stimulus intensity were 1.72 for ME1 and 1.15 for ME2 (effect sizes above *>* 1.2 are considered “very large”; Cohen, 1988), but only 0.45 for NRS, 0.48 for VAS, and 0.18 for gLMS, corresponding to small to medium effect sizes. Table A.4 to Table A.6 in the Appendix display all correlations among the different measures (and income).

**Table 2:** Reported experienced pain levels as measured by the different methods, split by condition (Low Pain/Analgesic vs. High Pain/Placebo). Low/High values are means, and *p*-values are for MWU tests. Entries in brackets give the SE or the *z* statistics.

|  | Electrical pain |  |  | Contact-heat pain |  |  | Analgesic |  |  |
| --- | --- | --- | --- | --- | --- | --- | --- | --- | --- |
|  | Low | High | p-value | Low | High | p-value | Analgesic | Placebo | p-value |
| NRS | 48.51<br>(3.24) | 59.35<br>(3.14) | 0.0234<br>(-2.261) | 39.24<br>(3.22) | 53.63<br>(3.18) | 0.0015<br>(-3.144) | 66.83<br>(2.39) | 56.32<br>(3.30) | 0.0133<br>(2.466) |
| VAS | 45.29<br>(3.12) | 56.52<br>(3.06) | 0.0145<br>(-2.436) | 40.17<br>(3.24) | 52.48<br>(3.07) | 0.0050<br>(-2.791) | 62.87<br>(2.34) | 55.61<br>(3.06) | 0.0750<br>(1.782) |
| gLMS | 55.27<br>(3.18) | 59.45<br>(3.06) | 0.3164<br>(-1.006) | 42.18<br>(3.18) | 52.51<br>(2.72) | 0.0114<br>(-2.520) | 63.58<br>(2.13) | 54.74<br>(2.41) | 0.0104<br>(2.550) |
| ME1 | 5.59<br>(0.35) | 16.43<br>(0.48) | $<0.0001$<br>(-9.044) | 4.53<br>(0.28) | 18.11<br>(0.72) | $<0.0001$<br>(-9.026) | 5.19<br>(0.52) | 20.66<br>(0.36) | $<0.0001$<br>(-8.971) |
| ME2 | 7.68<br>(1.05) | 16.94<br>(0.69) | $<0.0001$<br>(-7.198) | 6.42<br>(0.43) | 20.31<br>(0.71) | $<0.0001$<br>(-8.919) | 6.51<br>(0.69) | 19.53<br>(0.34) | $<0.0001$<br>(-8.328) |

For the contact-heat study, the differences were also significant in the expected direction for all methods (Table 2). Again, the separation between conditions was stronger for the new than for the standard methods: Effect sizes were 1.71 for ME1 and 1.68 for ME2 (“very large”). For the standard methods, effect sizes were 0.57 for NRS, 0.50 for VAS, and 0.45 for gLMS, corresponding to medium effect sizes.

For the analgesic study, the standard procedures failed to distinguish conditions in the expected direction, and even produced significant differences in the *unexpected* direction (Table 2). In contrast, the proposed methods resulted in highly-significant differences in the expected direction and separated the two conditions more strongly than the standard methods. Effect sizes were 1.84 for ME1 and 1.70 for ME2 (“very large”), while for NRS, VAS, and gLMS effect sizes were *−*0.49, *−*0.36, and *−*0.52, respectively, corresponding to medium effects in the wrong direction.

The failure of the standard scales in the analgesic study might be related to an expectation violation confound. 95.83% of participants in the analgesic condition wrongly thought they were assigned to the placebo, suggesting that they expected the analgesic to completely nullify the painful stimulus. When they felt pain, they concluded that they were in the placebo group and might have self-reported a high experienced pain. While the mechanism driving such an overcompensation (a possible nocebo effect; Enck et al., 2008) is unclear, the key observation for our purposes is that, when provided with a familiar (monetary) scale, participants’ choices were confound-free.

We remark that the Placebo condition of the analgesic study and the High condition of the contact-heat study were identical in terms of stimulus intensity. As expected, none of the measures were statistically significantly different across these two conditions (NRS *p* = 0.5695, VAS *p* = 0.4996, gLMS *p* = 0.4013, ME1 *p* = 0.4855, ME2 *p* = 0.4855). This speaks against a placebo effect in our third study and confirms the stability of the new measures.

### 3.2 Bayesian factor analysis

In summary, in all three studies, the data confirm that the new methods differentiate the perception of different levels of nociceptive input across subjects more strongly than the standard methods. We then assessed the differentiation strength of the various methods for pain measurement with a Bayesian factor analysis, which allows to quantify the probability with which a particular model is better at explaining the data compared to others. We used uninformed priors and set ME1 as the base category. For ease of comparison, all reported Bayesian factors refer to how much more likely ME1 is to explain the data than the alternative method considered in the comparison.

Log-Bayesian factors provided “decisive evidence” (Jeffreys, 1998) in all three studies that ME1 and ME2 were better predictors than the standard procedures (comparing ME1 with NRS, electrical pain: 64.63, contact heat: 61.63, analgesic: 62.76; ME1 with VAS, electrical pain: 64.22, contact heat: 62.75, analgesic: 64.18; ME1 with gLMS, electrical pain: 67.00, contact heat: 63.41, analgesic: 61.93). ME1 and ME2 themselves were comparable (log-Bayesian factors of ME1 with base ME2 *−*0.06 for electrical pain, 0.04 for contact heat, and 0.00 for analgesic).

### 3.3 Probit regressions

We also performed probit regressions on the probability of a participant being allocated to the High pain/Placebo condition as a function of the different measurements and demographic factors. The analyses, reported in Tables 3 to 5, confirmed the previous results, even when controlling for gender and income (results are unchanged when also controlling for valuation of money). The regressions do not include ME2 because it is highly correlated with ME1, but we find qualitatively the same results (omitted for the sake of brevity) when using ME2 instead of ME1.

**Table 3:** Probit regressions to predict whether a participant is in the High Pain condition for study 1 (electrical pain). Model 7 excludes people reporting no income. Standard errors are reported in parentheses. ^*\**^ *p <* 0.1, ^*\*\**^ *p <* 0.05, ^*\*\*\**^ *p <* 0.01.

| High Pain | Model 1 | Model 2 | Model 3 | Model 4 | Model 5 | Model 6 | Model 7 |
| --- | --- | --- | --- | --- | --- | --- | --- |
| NRS | 0.012**<br>(0.005) |  |  |  | -0.113<br>(0.077) | -0.149<br>(0.101) | -0.161<br>(0.164) |
| VAS |  | 0.013**<br>(0.005) |  |  | 0.144*<br>(0.081) | 0.159<br>(0.100) | 0.168<br>(0.163) |
| gLMS |  |  | 0.005<br>(0.005) |  | -0.011<br>(0.032) | -0.029<br>(0.036) | -0.029<br>(0.035) |
| ME1 |  |  |  | 0.600***<br>(0.139) | 0.983***<br>(0.367) | 0.986***<br>(0.379) | 0.966**<br>(0.414) |
| Female | -0.083<br>(0.249) | -0.073<br>(0.250) | -0.154<br>(0.244) | -0.737<br>(0.666) | -2.098<br>(1.388) | -2.480<br>(1.535) | -2.576<br>(1.716) |
| Income | -0.000<br>(0.000) | -0.000<br>(0.000) | -0.000<br>(0.000) | 0.000<br>(0.000) | 0.000<br>(0.000) |  | 0.000<br>(0.000) |
| Constant | -0.494<br>(0.354) | -0.531<br>(0.345) | -0.113<br>(0.352) | -6.667***<br>(1.659) | -10.534**<br>(5.012) | -8.322**<br>(4.127) | -7.969**<br>(4.007) |
| N | 109 | 109 | 109 | 109 | 109 | 109 | 81 |
| Pseudo $R^2$ | 0.049 | 0.055 | 0.022 | 0.852 | 0.902 | 0.885 | 0.879 |

**Table 4:** Probit regressions to predict whether a participant is in the High Pain condition for study 2 (contact-heat pain). Model 7 excludes people reporting no income. Standard errors are reported in parentheses. ^*\**^ *p <* 0.1, ^*\*\**^ *p <* 0.05, ^*\*\*\**^ *p <* 0.01.

| High Pain | Model 1 | Model 2 | Model 3 | Model 4 | Model 5 | Model 6 | Model 7 |
| --- | --- | --- | --- | --- | --- | --- | --- |
| NRS | 0.016***<br>(0.006) |  |  |  | 0.048<br>(0.040) | 0.045<br>(0.040) | 0.012<br>(0.038) |
| VAS |  | 0.014**<br>(0.006) |  |  | -0.032<br>(0.046) | -0.031<br>(0.046) | 0.034<br>(0.053) |
| gLMS |  |  | 0.014**<br>(0.006) |  | -0.005<br>(0.017) | -0.006<br>(0.017) | -0.048<br>(0.037) |
| ME1 |  |  |  | 0.343***<br>(0.061) | 0.379***<br>(0.080) | 0.377***<br>(0.076) | 0.353***<br>(0.117) |
| Female | -0.272<br>(0.271) | -0.223<br>(0.268) | -0.205<br>(0.265) | -0.681<br>(0.557) | -0.943<br>(0.668) | -0.882<br>(0.649) | -0.975<br>(0.877) |
| Income | 0.000<br>(0.000) | 0.000<br>(0.000) | 0.000<br>(0.000) | 0.000<br>(0.000) | 0.000<br>(0.000) |  | 0.000<br>(0.000) |
| Constant | -0.673**<br>(0.275) | -0.590**<br>(0.276) | -0.605**<br>(0.292) | -3.099***<br>(0.529) | -3.760***<br>(0.953) | -3.593***<br>(0.894) | -2.859**<br>(1.391) |
| N | 106 | 106 | 106 | 106 | 106 | 106 | 44 |
| Pseudo $R^2$ | 0.066 | 0.050 | 0.045 | 0.797 | 0.816 | 0.815 | 0.756 |

**Table 5:** Probit regressions to predict whether a participant is in the High Pain condition for study 3 (contact-heat and analgesic pain). Model 7 excludes people reporting no income. Standard errors are reported in parentheses. ^*\**^ *p <* 0.1, ^*\*\**^ *p <* 0.05, ^*\*\*\**^ *p <* 0.01.

| Placebo | Model 1 | Model 2 | Model 3 | Model 4 | Model 5 | Model 6 | Model 7 |
| --- | --- | --- | --- | --- | --- | --- | --- |
| NRS | -0.015**<br>(0.006) |  |  |  | -0.068<br>(0.055) | -0.067<br>(0.053) | -0.059<br>(0.054) |
| VAS |  | -0.011*<br>(0.006) |  |  | 0.052<br>(0.069) | 0.050<br>(0.066) | 0.029<br>(0.071) |
| gLMS |  |  | -0.021***<br>(0.008) |  | -0.043<br>(0.038) | -0.045<br>(0.039) | -0.064<br>(0.059) |
| ME1 |  |  |  | 0.324***<br>(0.081) | 0.369***<br>(0.104) | 0.375***<br>(0.109) | 0.405**<br>(0.185) |
| Female | 0.101<br>(0.263) | 0.070<br>(0.261) | 0.167<br>(0.266) | 0.229<br>(0.588) | 0.347<br>(0.790) | 0.311<br>(0.789) | 0.074<br>(0.860) |
| Income | -0.000<br>(0.000) | -0.000<br>(0.000) | -0.000<br>(0.000) | 0.000<br>(0.000) | 0.000<br>(0.000) |  | 0.000<br>(0.000) |
| Constant | 0.900**<br>(0.425) | 0.646<br>(0.432) | 1.185**<br>(0.497) | -4.973***<br>(1.595) | -1.592<br>(1.985) | -1.407<br>(1.893) | 0.257<br>(2.646) |
| N | 104 | 104 | 104 | 104 | 104 | 104 | 76 |
| Pseudo $R^2$ | 0.047 | 0.024 | 0.055 | 0.849 | 0.892 | 0.892 | 0.880 |

Specifically, models 1–3 show that NRS, VAS, and gLMS were mostly predictive of subjects’ assignment both for the electrical pain study (NRS, *t*(3) = 2.22, odd-ratio effect size 1.019; VAS, *t*(3) = 2.42, effect size 1.034; gLMS, *t*(3) = 0.97 (n.s.), effect size 1.018) and the contact-heat pain study (NRS, *t*(3) = 2.91, effect size 1.026; VAS, *t*(3) = 2.50, effect size 1.037; gLMS, *t*(3) = 2.37, effect size 1.052). For the analgesic study, the three standard measures incorrectly predicted subjects’ assignment, i.e. coefficients were significant in the wrong direction (NRS, *t*(3) = *−*2.49, effect size 1.024; VAS, *t*(3) = *−*1.74, effect size 1.029; gLMS, *t*(3) = *−*2.67, effect size 1.079). For all studies, ME1 was the best predictor of higher acute pain in the High pain/Placebo conditions (Model 4: Study 1, *t*(3) = 4.32, effect size 15.800; Study 2, *t*(3) = 5.62, effect size 4.844; Study 3, *t*(3) = 4.00, effect size 4.439).

Moreover, as shown by Model 5, when all measures are included as predictors, ME1 remains the only significant variable (Study 1, *t*(6) = 2.68, effect size 245.869; Study 2, *t*(6) = 4.73, effect size 8.351; Study 3, *t*(6) = 3.53, effect size 7.896). Moreover, ME1 explains a substantial part of the variance in the data, with all pseudo-*R*^2^s above 0.816 in the joint models and above 0.797 when regressed alone, while the *R*^2^s of the other methods is below 0.1. Tables 3 to 5 also confirm that income and gender did not add any explanatory power for whether a participant was in the High pain/Placebo condition.

## 4 Discussion

In this work we show that standard economic methods can improve the measurement of subjective, individual experiences. In particular, we focus on the elicitation of experienced (acute) pain as a socially and economically relevant case. With this aim, we proposed and validated a new method to measure the experienced intensity of acute pain across individuals. This method proved to be overwhelmingly more accurate and sensitive than standard, established methods for experimentally-induced acute pain which are used in medical, clinical and research applications. The findings hold for different pain modalities and for an analgesic treatment where all participants received an isointense nociceptive stimulus, i.e., a stimulus with the same physical intensity.

Our work contributes to the ongoing debate on the use of subjective, numerical scales to measure human feelings. Such scales are often argued to lack scientific foundations (Bond and Lang, 2019), but recent research has suggested that they do have predictive power (Kaiser and Oswald, 2022). We build a bridge between both views by showing that switching to a less-subjective scale (if at all possible) can greatly improve the reliability of measurements.

In the case of experienced pain intensity, the mechanism that enables the new measurement method to excel might indeed be rooted in its well-calibrated scale: money. Money-free standard methods ask people for numerical estimations of their experiences without a benchmark for the numerical scale, which might lead them to choose salient numbers or intermediate values. A similar argument can be made for graphical procedures involving marks on abstract lines. Moreover, it is conceivable that a given number, say 3, or a given distance on a segment, is not comparable across different people, especially when it refers to an abstract and personal construct such as pain. Monetary amounts provide interpersonally comparable references and thereby reduce the intrinsic subjectivity in pain measurements.

Our results may also have implications for the measurement of perceived pain in clinical practice. Our third study emulated a standard procedure to evaluate the effectiveness of an analgesic treatment (Huskisson, 1974; Chow et al., 2009), i.e., a double-blind randomized controlled trial, where pain is measured in a controlled environment and compared between the two different arms of the treatment. The new method could in principle be integrated into clinical trials focused on random parallel-assignment, e.g. those investigating the effectiveness of analgesics. The use of monetary incentives is unproblematic for those applications, as it is already common practice to reward participation in them (Wallenstein et al., 1980).

The limitations of our studies are as follows. First, we focused on experimentally-induced pain on healthy subjects. Before eventual clinical applications, research should validate the proposed protocol in different populations of patients and consider implementations closer to clinical pain. Second, our studies employed only two different (moderate) levels of nociceptive input, since our objective was to test how well the different measures could distinguish them between-subjects. Future studies could vary stimulus intensity along a more extensive and fine-grained spectrum including milder and more extreme pain levels. Third, we focused on evoked pain, which is different from spontaneous clinical pain. Pain is known to be multidimensional (Clark et al., 2002), and our method targets its motivational aspects (related to pain affect and pain avoidance). Future research should clarify which aspects of pain are targeted by specific methods. Fourth, as a benchmark, we focused on the most prominent and commonly-used scales in practice (NRS, VAS, and gLMS). Further research could compare to other existing scales, as, e.g., the multidimensional affect and pain survey (MAPS) questionnaire of Clark et al. (2002). Fifth, for clinical applications beyond randomized controlled trials, monetary incentives might need to be replaced with hypothetical questions, requiring further validation. This might be the case, for instance, for studies on analgesic efficiency after clinical procedures, which currently often rely on scales such as NRS (Singla et al., 2017; Viscusi, 2022). Further, individual differences in wealth and income might play a role in the precision and between-subject comparability of the new method, requiring specific controls. We controlled for these possible factors in our studies and showed that they did not play a role in our results. However, the vast majority of our participants were students, many of whom reported zero or low income. Hence, subsequent work should study this issue in different populations.

In conclusion, our studies support the view that standard pain measurements, which are a prominent example of self-reported numerical scales, are noisy and exhibit limited correlation (Huskisson, 1974; Breivik et al., 2008; Phillips, 2009; Von Korff et al., 2020), making it difficult to evaluate whether pain has been accurately measured and adequately relieved. Our new method, based on standard economic elicitation procedures, provides a complementary approach to reliably measure experienced pain across individuals and could open the door to future research improving pain measurement and management. More generally, our contribution shows that the reliability of subjective, numerical scales measuring human feelings can sometimes be greatly improved by switching the focus to instrumental quantities allowing for better interpersonal comparisons.

## Data Availability

De-identified participants data and the code for the analyses is publicly available in OSF

https://osf.io/jtv8p

## Appendix

### Supplementary Tables

**Table A.1:**
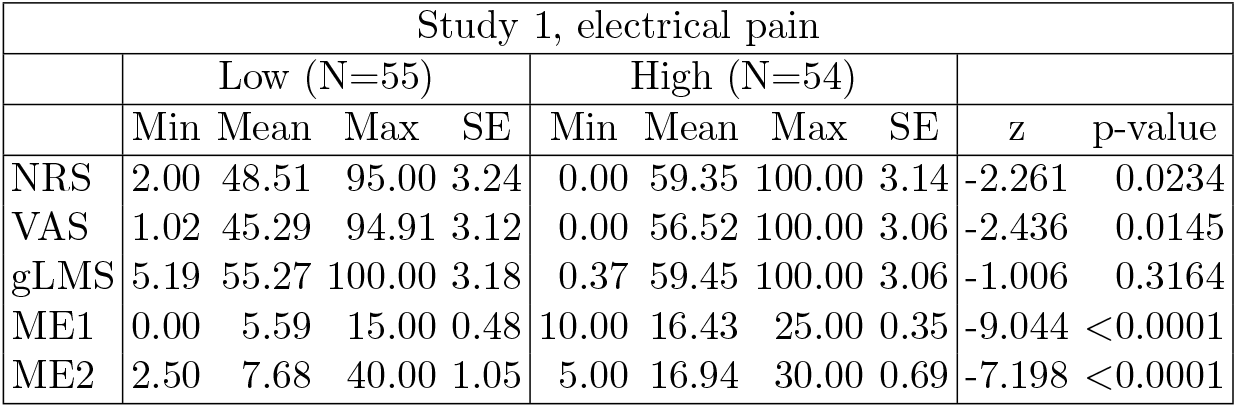
Reported experienced pain levels as measured by the different methods, split by condition (Low Pain vs. High Pain). Low/High values are min, means, max, standard errors, *z* statistics and *p*-values are for MWU tests.

**Table A.2:** Reported experienced pain levels as measured by the different methods, split by condition (Low Pain vs. High Pain). Low/High values are min, means, max, standard errors, *z* statistics and *p*-values are for MWU tests.

| Study 2, contact-heat pain |  |  |  |  |  |  |  |  |  |  |
| --- | --- | --- | --- | --- | --- | --- | --- | --- | --- | --- |
|  | Low (N=58) |  |  |  | High (N=57) |  |  |  |  |  |
|  | Min | Mean | Max | SE | Min | Mean | Max | SE | z | p-value |
| NRS | 5.00 | 39.24 | 100.00 | 3.22 | 15.00 | 53.63 | 100.00 | 3.18 | -3.144 | 0.0015 |
| VAS | 2.96 | 40.17 | 100.00 | 3.24 | 8.61 | 52.48 | 99.63 | 3.07 | -2.791 | 0.0050 |
| gLMS | 4.44 | 42.18 | 96.85 | 3.18 | 10.37 | 52.51 | 99.74 | 2.72 | -2.520 | 0.0114 |
| ME1 | 2.50 | 4.53 | 10.00 | 0.28 | 0.00 | 18.11 | 30.00 | 0.72 | -9.026 | <0.0001 |
| ME2 | 2.50 | 6.42 | 20.00 | 0.43 | 5.00 | 20.31 | 40.00 | 0.71 | -8.919 | <0.0001 |

**Table A.3:**
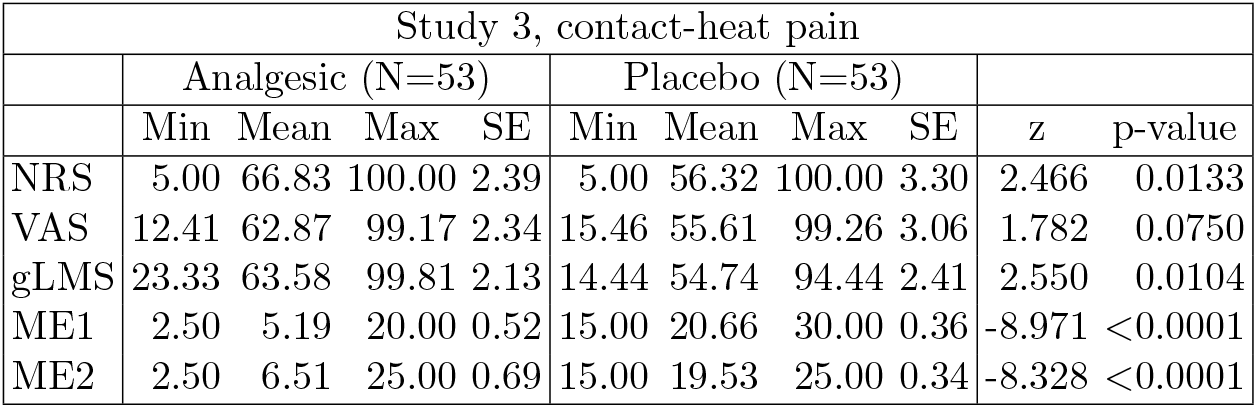
Reported experienced pain levels as measured by the different methods, split by condition (Low Pain vs. High Pain). Low/High values are min, means, max, standard errors, *z* statistics and *p*-values are for MWU tests.

**Table A.4:** Study 1: Correlation among the different measures of pain and income. p-values in parenthesis.

|  | ME1 | ME2 | NRS | VAS | gLMS | Income |
| --- | --- | --- | --- | --- | --- | --- |
| ME1 | 1.000 |  |  |  |  |  |
|  | - |  |  |  |  |  |
| ME2 | 0.604<br>(0.000) | 1.000 |  |  |  |  |
|  |  | - |  |  |  |  |
| NRS | 0.177<br>(0.065) | 0.232<br>(0.016) | 1.000 |  |  |  |
|  |  |  | - |  |  |  |
| VAS | 0.172<br>(0.073) | 0.183<br>(0.057) | 0.969<br>(0.000) | 1.000 |  |  |
|  |  |  |  | - |  |  |
| gLMS | 0.134<br>(0.164) | 0.129<br>(0.182) | -0.424<br>(0.000) | -0.408<br>(0.000) | 1.000 |  |
|  |  |  |  |  | - |  |
| Income | -0.214<br>(0.025) | -0.196<br>(0.042) | -0.071<br>(0.461) | -0.038<br>(0.694) | -0.004<br>(0.971) | 1.000 |
|  |  |  |  |  |  | - |

**Table A.5:** Study 2: Correlation among the different measures of pain and income. p-values in parenthesis.

|  | ME1 | ME2 | NRS | VAS | gLMS | Income |
| --- | --- | --- | --- | --- | --- | --- |
| ME1 | 1.000 |  |  |  |  |  |
|  | - |  |  |  |  |  |
| ME2 | 0.833<br>(0.000) | 1.000 |  |  |  |  |
|  |  | - |  |  |  |  |
| NRS | 0.315<br>(0.001) | 0.370<br>(0.000) | 1.000 |  |  |  |
|  |  |  | - |  |  |  |
| VAS | 0.289<br>(0.002) | 0.339<br>(0.000) | 0.959<br>(0.000) | 1.000 |  |  |
|  |  |  |  | - |  |  |
| gLMS | 0.268<br>(0.004) | 0.273<br>(0.003) | 0.820<br>(0.000) | 0.826<br>(0.000) | 1.000 |  |
|  |  |  |  |  | - |  |
| Income | 0.001<br>(0.996) | -0.019<br>(0.843) | -0.016<br>(0.865) | -0.016<br>(0.870) | 0.011<br>(0.907) | 1.000 |
|  |  |  |  |  |  | - |

**Table A.6:** Study 3: Correlation among the different measures of pain and income. p-values in parenthesis.

|  | ME1 | ME2 | NRS | VAS | gLMS | Income |
| --- | --- | --- | --- | --- | --- | --- |
| ME1 | 1.000 |  |  |  |  |  |
|  | - |  |  |  |  |  |
| ME2 | 0.913<br>(0.000) | 1.000 |  |  |  |  |
|  |  | - |  |  |  |  |
| NRS | -0.149<br>(0.128) | -0.136<br>(0.165) | 1.000 |  |  |  |
|  |  |  | - |  |  |  |
| VAS | -0.090<br>(0.358) | -0.079<br>(0.422) | 0.921<br>(0.000) | 1.000 |  |  |
|  |  |  |  | - |  |  |
| gLMS | -0.222<br>(0.022) | -0.227<br>(0.019) | 0.349<br>(0.000) | 0.367<br>(0.000) | 1.000 |  |
|  |  |  |  |  | - |  |
| Income | -0.090<br>(0.361) | -0.034<br>(0.732) | 0.073<br>(0.462) | 0.115<br>(0.242) | -0.011<br>(0.915) | 1.000 |
|  |  |  |  |  |  | - |

